# Evaluating Clinical Concept Extraction and Evidence-Bounded Terminology Linking: Multisite Model Comparison and Pilot Ablation Study

**DOI:** 10.64898/2026.08.20.26360740

**Authors:** Yibo Chen, Mihail Popescu

## Abstract

**Background:** Clinical terminology pipelines must first extract candidate spans from narrative notes and then determine whether those spans map to existing concepts or warrant further review. Evaluation is difficult because span boundaries vary between annotators and because downstream decisions depend on the terminology evidence retrieved for each span.

**Objective:** We evaluated clinical concept extraction, terminology linking across controlled evidence conditions, and ontology-extension triage for terms that remained unmatched after initial terminology screening.

**Methods:** We conducted 3 complementary pilot evaluations that used distinct units of analysis and were analyzed separately. Study 1 compared 5 automated extraction pipelines and a union-merge analysis with 2 human annotation sets in 66 deidentified clinical notes from 3 health systems. Agreement was evaluated by exact string matching and BGE-large-en-v1.5 embedding matching. Study 2 evaluated 56 clinical spans—28 with reference Unified Medical Language System (UMLS) concepts and 28 adjudicated as unsuitable for ontology extension—under complete retrieval, matched-concept masking, and large language model (LLM)-only inference, yielding 168 span-condition outputs. The graph retrieval pipeline used BGE-large-en-v1.5 embeddings, and the decision model was Gemma 3 27B. Study 3 applied full vector retrieval to 84 terms previously not matched in either UMLS or BioPortal.

**Results:** In Study 1, interannotator exact-match F1 was 0.29 and embedding-match F1 was 0.75. Automated exact-match F1 scores ranged from 0.07 to 0.17; embedding-match F1 was highest for MedGemma (0.55), followed by Gemma (0.53), `sci_md` and SciBERT (each 0.43), and Llama 3.3 (0.32). In Study 2, complete retrieval returned a reference-matched link for 28/28 known-concept spans (100%; 95% CI, 87.9%-100%). Masking assigned `POSSIBLE_CANDIDATES` to all 28; LLM-only inference assigned `POSSIBLE_CANDIDATES` to 25/28 (89.3%) and `LINKED` to 3/28 (10.7%). Across the 3 evidence conditions, the same 12/28 unsuitable-extension spans were classified as `NOT_MEANINGFUL` (42.9%) and the same 16/28 as `POSSIBLE_CANDIDATES` (57.1%). In Study 3, the pipeline assigned `PLAUSIBLE_EXISTING_CONCEPT` to all 84 terms, none was flagged for extension, and top-candidate similarity averaged 0.914 (SD 0.027); extension status was not independently adjudicated.

**Conclusions:** Measured extraction performance varied substantially by matching definition, whereas exact-link decisions varied with the availability of matched terminology evidence. In the follow-up sample, initial nonmatching did not establish ontology novelty: after semantic retrieval, the pipeline classified all 84 terms as plausible existing concepts and proposed none for extension. These findings support separate evaluation of extraction, retrieval, evidence-grounded linking, and extension candidacy.

## Introduction

### Background

Clinical information extraction begins with identifying clinically relevant spans in narrative text. Named entity recognition (NER) performance is commonly measured by exact span agreement, but exact scoring penalizes boundary variation even when annotators or systems identify semantically similar concepts. Clinical evaluations therefore benefit from distinguishing exact boundary agreement from broader semantic correspondence and from interpreting automated performance in the context of human annotation variability.

Clinical concept normalization maps mentions in narrative text to standardized concepts, commonly represented by Unified Medical Language System (UMLS) Concept Unique Identifiers (CUIs) [1,2]. Reliable normalization supports semantic interoperability, cohort construction, phenotyping, clinical search, decision support, and secondary reuse of electronic health record data. The task remains difficult because clinical notes contain abbreviations, misspellings, local expressions, compositional phrases, and observations whose interpretation depends on both clinical context and the terminology available to the system [2].

Most contemporary normalization systems separate candidate generation from candidate selection [3–6]. A sparse, dense, or hybrid retrieval component first identifies plausible terminology concepts, after which a ranking or decision component chooses a concept or declines to link. Biomedical synonym representation learning and UMLS-aware candidate generation have improved this process [3–6], and recent work has combined semantic retrieval with LLM prompting for normalization [7]. This decomposition nevertheless creates a structural dependency: the downstream model cannot select a reference concept that the candidate generator did not retrieve [6].

Retrieval-augmented generation (RAG) conditions model output on external evidence [8]. Biomedical knowledge graph approaches similarly provide graph-derived context for reasoning and provenance [9,10]. Graph retrieval can be especially useful for terminology work because concepts, synonyms, semantic types, and relations can be presented as inspectable evidence. At the same time, retrieval introduces its own failure modes. Candidate context may be incomplete, noisy, or internally inconsistent, and an LLM may rely on parametric familiarity instead of the supplied evidence. Component-level evaluation is therefore needed to distinguish retrieval behavior from downstream decision behavior [11,12]. Here, graph RAG refers to retrieving candidate concepts and graph-derived context for an LLM decision layer rather than to corpus-level community summarization [13].

### Clinical Normalization Under Incomplete Retrieval

Incomplete retrieval creates a consequential ambiguity. If the correct concept is absent from a candidate set, the model has not established that no corresponding terminology concept exists. It has established only that the visible evidence does not support one. An exact link asserted under those circumstances may be semantically plausible but lacks visible provenance. Conversely, interpreting every retrieval failure as evidence of novelty would create inappropriate extension proposals for concepts already represented elsewhere in the ontology.

An evidence-bounded workflow therefore benefits from an explicit deferral state. Selective prediction formalizes a reject option when a model lacks sufficient evidence [14]. CUI-less normalization similarly recognizes that some spans should remain unlinked [15]. Ontology-extension triage adds another distinction: some unlinked spans may be potentially novel concepts, whereas others are clinically meaningful events, instructions, compositional phrases, or documentation fragments that are not suitable as independent ontology nodes. These cases require different downstream actions even though neither should receive an unsupported exact link.

Study 2 used 3 output labels. `LINKED` indicated an exact terminology link supported by visible evidence. `POSSIBLE_CANDIDATES` indicated a clinically meaningful span retained for additional retrieval or review because the visible evidence did not support an exact link. The system used `NOT_MEANINGFUL` for spans adjudicated as unsuitable for ontology extension; here, this label denotes unsuitability as an independent extension candidate, not absence of clinical meaning. Study 3 used a different structured output consisting of a triage label, including `PLAUSIBLE_EXISTING_CONCEPT`, and a separate binary extension flag.

### Study Objective

We conducted 3 complementary evaluations of a clinical terminology workflow. Study 1 compared automated clinical concept extraction with double human annotation in 66 deidentified notes from 3 health systems. Study 2 was a paired ablation examining how retrieval completeness affected the terminology-linking decision layer. Study 3 characterized a later follow-up sample of clinical terms that had not received initial matches in UMLS or BioPortal and were submitted to full vector retrieval and extension triage. The evaluations used distinct units, reference standards, and outcomes and were analyzed separately.

The studies addressed 5 questions: how exact and embedding-based matching changed measured NER agreement; whether the decision layer selected a known UMLS concept when its matched node was available; whether withholding that node induced deferral; whether retrieved evidence improved rejection of unsuitable extension candidates; and how the pipeline triaged a discipline-balanced follow-up sample of initially unmatched terms. We expected complete evidence to support exact linking and incomplete evidence to increase deferral. We treated initial nonmatching as candidate-generation evidence, not proof of ontology novelty.

## Methods

### Study Design

This work was conducted within the Creating AI-Enabled All-Health Team Data Fabric (CAIDF) project, a 5-institution collaboration developing multidisciplinary data and discharge-summary methods for neonatal intensive care unit and adult fall-related hospitalization cohorts. The present studies used records from the 3 data-contributing health systems and evaluated terminology-processing components rather than discharge-summary performance.

The project evaluated 3 complementary components of clinical terminology processing: extraction of candidate spans, evidence-sensitive terminology linking, and follow-up extension triage. The evaluations used notes, spans, and terms as distinct units of analysis and were analyzed separately. Record-level linkage across studies was unavailable, and no end-to-end performance estimate was calculated. Figure 1 summarizes the analytical structure.

**Figure 1.**
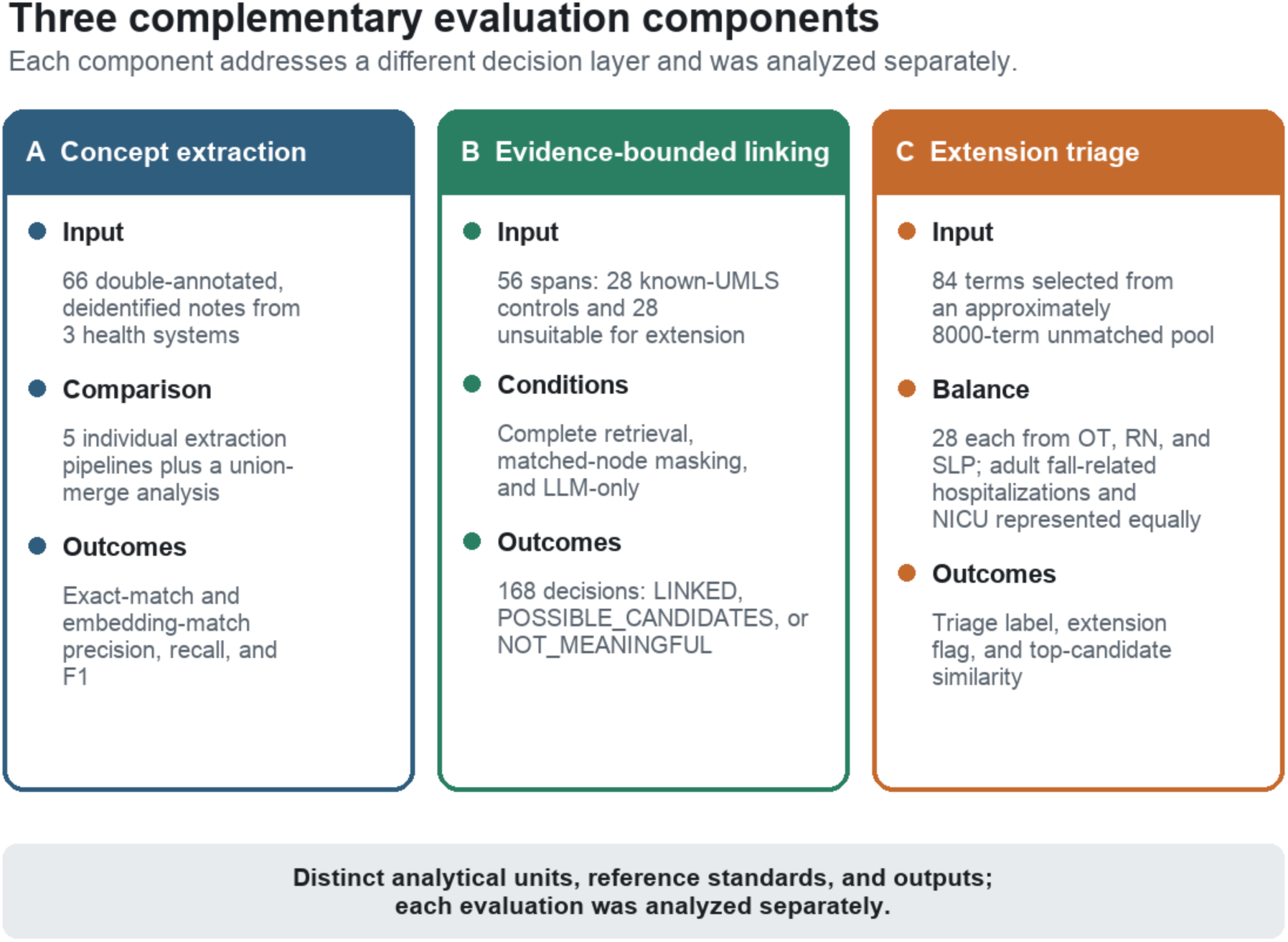
Analytical structure of the 3 complementary component evaluations. Study 1 assessed clinical concept extraction in 66 double-annotated notes from 3 health systems. Study 2 evaluated 56 spans under 3 parallel evidence conditions, producing 168 span-condition decisions. Study 3 characterized 84 discipline-balanced terms selected from an approximately 8000-term pool previously screened as unmatched to UMLS or BioPortal. The studies used distinct analytical units, reference standards, and output taxonomies and were analyzed separately. LLM: large language model; NICU: neonatal intensive care unit; OT: occupational therapy; RN: nursing; SLP: speech-language pathology; UMLS: Unified Medical Language System.

Study 1 was a multisite clinical concept extraction comparison. Study 2 was an exploratory, within-item pilot ablation. The same 56 clinical spans were evaluated under 3 evidence conditions, yielding 168 span-condition evaluations. Each span served as its own control. The experimental manipulation changed the evidence supplied to the decision layer while retaining the same prompt template, output schema, and decision labels.

Study 2 isolated evidence-sensitive decision behavior rather than estimating end-to-end clinical normalization performance. The complete-evidence condition deliberately included the reference concept for known-UMLS spans and served as a manipulation check of whether the decision layer could use decisive evidence. The masked condition simulated a retrieval miss by removing that evidence. The retrieval-free condition examined whether the LLM would make exact-link decisions from parametric familiarity alone.

Study 3 was a single-condition follow-up of terms that remained unmatched after initial screening. From an approximately 8000-term pool previously screened as unmatched to UMLS or BioPortal, the analysis set included 84 terms: 28 each from the occupational therapy (OT), nursing (RN), and speech-language pathology (SLP) domains. Within each discipline, 14 terms came from the adult fall-related hospitalization cohort and 14 from the neonatal intensive care unit (NICU) cohort. Full vector retrieval returned 10 visible candidates for each term. Study 3 was analyzed separately because its sample, output taxonomy, and design differed from Study 2.

### Clinical Concept Extraction Evaluation

Study 1 used a 66-note analytic subset of the CAIDF double-annotated corpus from University of Missouri Health Care, University of Illinois Chicago, and the University of Iowa. The notes represented adult fall-related hospitalizations and neonatal intensive care contexts and included multidisciplinary documentation. The parent corpus was curated by discipline-specific clinical teams, which selected clinically information-rich notes using cohort- and discipline-specific eligibility criteria and an adapted Modified Physician Documentation Quality Instrument. Selection disagreements were resolved through consensus. Clinicians annotated notes in ATLAS.ti using a shared framework anchored in 5 cTAKES concept categories and supplemented with discipline-specific codes. The 66 notes in Study 1 had 2 independent human annotation sets. Annotators labeled general clinical concepts—including medications, symptoms, disorders, procedures, and anatomical sites—as well as discipline-specific concepts.

Five individual extraction pipelines were evaluated: the scispaCy `sci_md` configuration [16], a SciBERT-based configuration [17], and 3 LLM pipelines (Llama 3.3 [18], Gemma, and MedGemma [19]). Study 2 separately used Gemma 3 27B as the terminology-linking decision model. A union-merge analysis was also evaluated.

Automated outputs were compared with the human annotation sets using exact string matching and embedding-based matching with BGE-large-en-v1.5 [20]. Precision, recall, and F1 were microaveraged across notes. Agreement between the 2 human annotation sets was calculated under the exact and embedding-based definitions to contextualize model-human agreement; it was not treated as a formal upper performance bound. Token-level human agreement was summarized with Cohen kappa and Krippendorff alpha.

### Graph Retrieval-Augmented Clinical Terminology Pipeline

Candidate retrieval was implemented in Neo4j using `VectorCypherRetriever` from the Neo4j GraphRAG package and locally stored BGE-large-en-v1.5 sentence embeddings [20]. Each input span was searched against concept-text and alias-text vector indexes. Alias hits were resolved to canonical concept nodes, duplicate concepts were merged, and candidates were ranked by vector similarity. Candidate concepts were expanded through undirected 1-hop `RELATED` relationships, and the resulting evidence was serialized as text for the generator. The complete-evidence condition presented 10 candidates to the decision layer. Similarity scores were used for ranking and were not interpreted as calibrated probabilities.

The decision layer used Gemma 3 27B with default generation settings [21]. Each input span was supplied with the retrieved context and a fixed instruction template; the model returned a structured decision and, when applicable, a selected terminology concept.

The decision policy assigned LINKED only when the visible candidate context supported an exact terminology match; POSSIBLE_CANDIDATES when the span was clinically interpretable but the evidence was insufficient for an exact link; and NOT_MEANINGFUL when the span was unsuitable as an independent ontology-extension candidate.

The instruction template and output taxonomy were held constant across the 3 Study 2 conditions. Each span was evaluated under all 3 conditions, and paired analyses used the span as the matching unit.

### Benchmark Construction and Reference Status

The pilot benchmark comprised 56 spans derived from adjudicated outputs of the clinical terminology pipeline. Spans originated from physician (MD), occupational therapy (OT), nursing (RN), and speech-language pathology (SLP) note domains. The benchmark contained 2 equal-sized reference subsets.

The first subset contained 28 spans with known UMLS representations and a corresponding matched graph node. These known-concept controls were used to evaluate whether the decision layer selected the reference concept when it was visible and deferred when it was not. The second subset contained 28 spans adjudicated as unsuitable for ontology extension. These spans were used to evaluate reference-concordant rejection as `NOT_MEANINGFUL`. Within this second subset, 7 spans came from MD notes, 11 from OT notes, 4 from RN notes, and 6 from SLP notes.

The known-UMLS subset served as a silver-standard control set based on project adjudication. Because the benchmark did not include an independently adjudicated extension-positive reference set, it evaluated exact-link commitment, deferral, and rejection but not end-to-end ontology-extension discovery.

The clinical span was the unit of analysis. Patient, encounter, and note identifiers were excluded; consequently, clustering at those levels could not be assessed. The pilot did not include duplicate independent adjudication or formal interrater-reliability assessment of the reference labels.

### Evidence Conditions

Table 1 summarizes the 3 evidence conditions.

**Table 1.** Evidence conditions used in the paired pilot ablation.

| Condition | Visible terminology evidence | Candidate context | Evaluation purpose |
| --- | --- | --- | --- |
| GraphRAG Full | Complete retrieved context; matched node visible for known-UMLS controls | 10 candidates per item | Decision behavior conditional on successful retrieval |
| GraphRAG Masked | Matched node and associated exact-match evidence removed | 4-9 alternative candidates for known controls | Decision behavior under deliberately incomplete retrieval |
| LLM-only | No retrieved terminology evidence | 0 candidates | Retrieval-free exact-link and deferral behavior |

In the complete retrieval condition, termed GraphRAG Full, the model received 10 visible candidates for each item. For known-UMLS spans, the context included the matched terminology node and its exact-match evidence. This condition measured the decision layer’s behavior conditional on successful candidate retrieval.

In the masked retrieval condition, termed GraphRAG Masked, the matched node and associated exact-match evidence were removed before inference. Known-UMLS spans retained alternative vector-neighbor candidates, with 4-9 candidates visible. Masking therefore changed both candidate identity and context size; the experiment evaluates incomplete evidence broadly and does not isolate removal of the matched node from the reduction in candidate count.

In the LLM-only condition, the evidence block was empty. This condition tested whether the model would make exact-link decisions without visible terminology support. Because the prompt and label definitions were unchanged, observed differences represent decision behavior under the 3 evidence settings.

For known-UMLS spans, masking removed the designated matched node and associated exact-match evidence. The analysis did not separately distinguish alias-, synonym-, or neighbor-level exclusions. No analogous reference node was designated for the unsuitable-extension subset; results for that subset are therefore interpreted descriptively rather than as an isolated node-removal experiment.

### Outcome Definitions

The primary outcome for the known-UMLS subset was a reference-matched `LINKED` decision. A `POSSIBLE_CANDIDATES` decision represented deferral when visible evidence was insufficient. A `NOT_MEANINGFUL` decision on a known concept would represent inappropriate rejection.

For spans adjudicated as unsuitable for extension, the primary outcome was concordance with the reference label `NOT_MEANINGFUL`. A `POSSIBLE_CANDIDATES` decision indicated retention for further review, whereas `LINKED` indicated a reference-discordant exact-link decision. Because the purpose-built benchmark was not prevalence representative, we report the reference-concordant rejection proportion rather than diagnostic specificity.

We also measured exact categorical agreement between each pair of evidence conditions. Qualitative review focused on spans whose decisions changed with evidence availability and on recurrent types of unsuitable spans that were retained for review.

For Study 3, outcomes were defined by the structured decision and extension-flag fields and were not mapped onto the 3-label Study 2 taxonomy. Study 3 therefore characterizes pipeline-level outputs rather than the behavior of the generator in isolation.

### Statistical Analysis

For Study 1, microaveraged precision, recall, and F1 were calculated under exact and embedding-based matching. System comparisons were descriptive; no uncertainty intervals or paired inferential comparisons were calculated. Interannotator concept- and token-level agreement characterized annotation variability.

For Study 2, categorical outcomes are reported as n/N (%) with 2-sided Wilson 95% CIs. For the known-UMLS subset, paired differences in binary `LINKED` status were evaluated with exact McNemar tests. Exact categorical agreement was calculated for each condition pair across all 56 spans, within the known-UMLS subset, and within the unsuitable-extension subset. Reference-concordant rejection was summarized descriptively by note discipline.

The analyses were exploratory and were not preregistered; no prospective sample-size calculation was performed. Discipline-specific results were summarized descriptively because subgroup sizes were small and uneven. All hypothesis tests were 2-sided, and unadjusted *P* values are reported.

For Study 3, we summarized term distributions by discipline and clinical setting, structured triage and extension decisions, and top-candidate vector similarity using the mean, SD, median, IQR, and range. Similarity was treated as an uncalibrated retrieval-ranking score rather than a probability. Extension accuracy was not calculated because independently adjudicated reference labels for extension candidacy were unavailable.

Wilson confidence intervals were calculated from aggregate counts, and exact McNemar tests were performed on paired condition outcomes, using Python 3.12.6 and statsmodels 0.14.0.

### Ethical Considerations

This study was conducted as a secondary analysis within the Creating AI-Enabled All-Health Team Data Fabric project. The parent project was reviewed and approved by the University of Illinois Chicago Institutional Review Board (protocol 2024-0553), and each of the 3 data-contributing institutions obtained the required regulatory approvals before project initiation. Institutional review board approvals, data use agreements, and institutional data-governance procedures were completed before data acquisition, and investigators accessed and analyzed project data under these approvals and agreements.

The present analyses used deidentified clinical notes and derived spans and terms without patient, encounter, or note identifiers. Before transfer to the secure University of Illinois Chicago computing enclave, University of Missouri Health Care deidentified its notes using an institutional anonymization pipeline; University of Illinois Chicago and the University of Iowa used TackleAI under separate Business Associate Agreements. Following quality assurance, deidentified data were transferred to the secure enclave, which met National Institute of Standards and Technology Special Publication 800-171 requirements and required institutional credentials and 2-factor authentication. The illustrative spans reported in this manuscript were drawn from the deidentified analytic materials and contain no direct identifiers. The reported analyses involved no direct participant contact or study-specific compensation.

## Results

### Clinical Concept Extraction and Human Agreement

Agreement varied substantially with the matching definition (Figure 2). Interannotator F1 was 0.29 under exact matching and 0.75 under embedding-based matching. Among individual automated systems, exact-match F1 ranged from 0.07 for Llama 3.3 to 0.17 for MedGemma. Under embedding-based matching, MedGemma had the highest observed automated-system F1 at 0.55, followed by Gemma at 0.53, `sci_md` and SciBERT at 0.43 each, and Llama 3.3 at 0.32. No inferential comparisons among extraction systems were performed.

**Figure 2.**
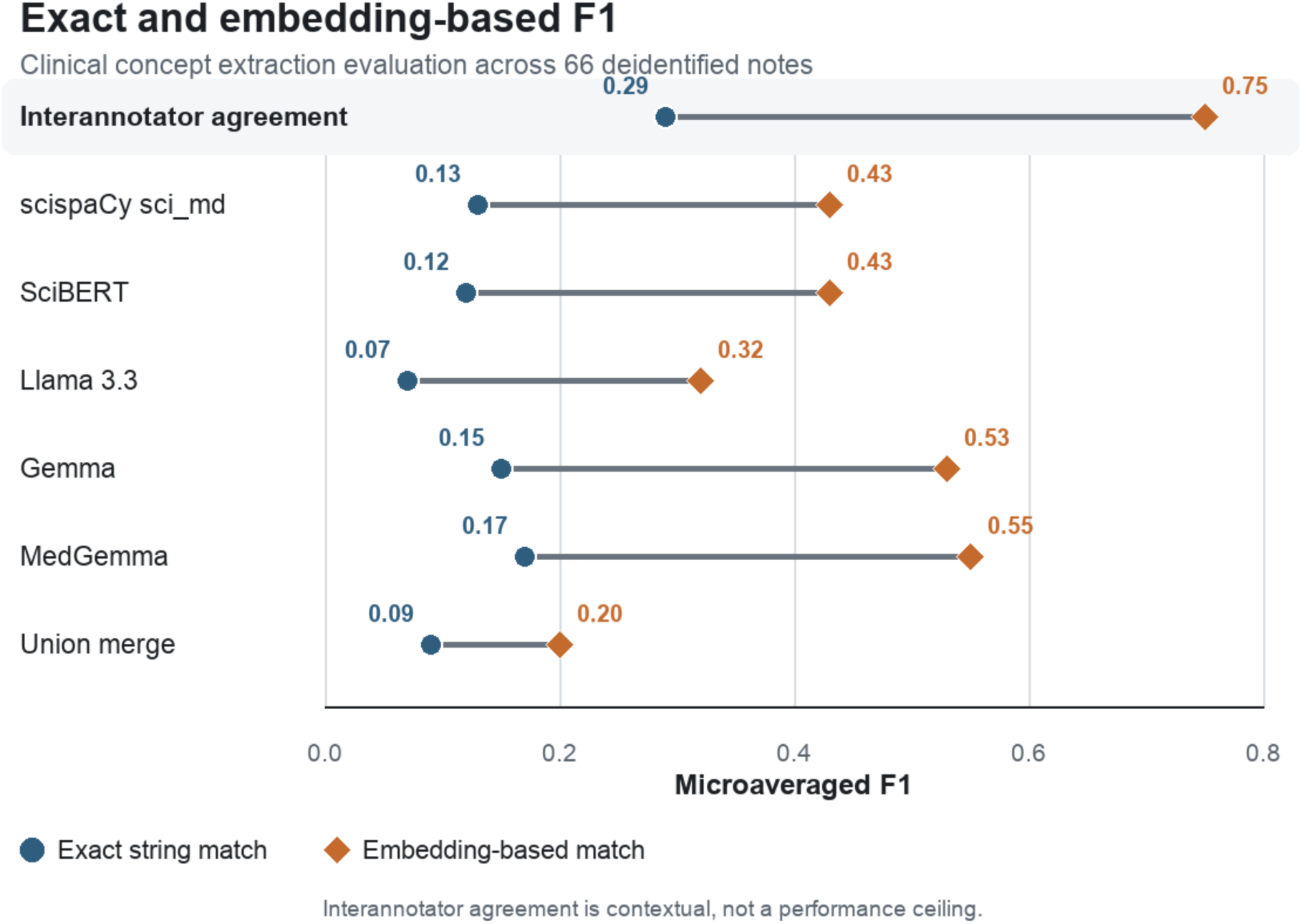
Exact- and embedding-match F1 scores for interannotator agreement and automated or fused extraction outputs across 66 deidentified clinical notes. Scores were microaveraged across notes. Circles indicate exact string matching, and diamonds indicate embedding-based matching using BGE-large-en-v1.5; numeric labels provide the exact values. Interannotator agreement contextualizes annotation variability and is not a formal upper performance bound. No inferential comparisons among extraction systems were performed.

Union merging emphasized recall. Under exact matching, it produced precision of 0.05, recall of 0.41, and F1 of 0.09. Under embedding-based matching, precision was 0.11, recall was 0.94, and F1 was 0.20. At the token level, Cohen kappa was 0.39 and Krippendorff alpha was 0.33 across 8665 token units.

### Evidence-Completeness Benchmark Characteristics

All 56 spans were evaluated under all 3 conditions, yielding 168 categorical decisions. The benchmark included 28 known-UMLS controls and 28 spans adjudicated as unsuitable for ontology extension. The complete retrieval condition provided 10 candidates per item. For known controls, masking removed the matched node and reduced the visible set to 4-9 alternative candidates. The LLM-only condition provided no candidate evidence.

### Decisions Across Evidence Conditions

Paired decision transitions across the 3 evidence conditions are shown in Figure 3.

**Figure 3.**
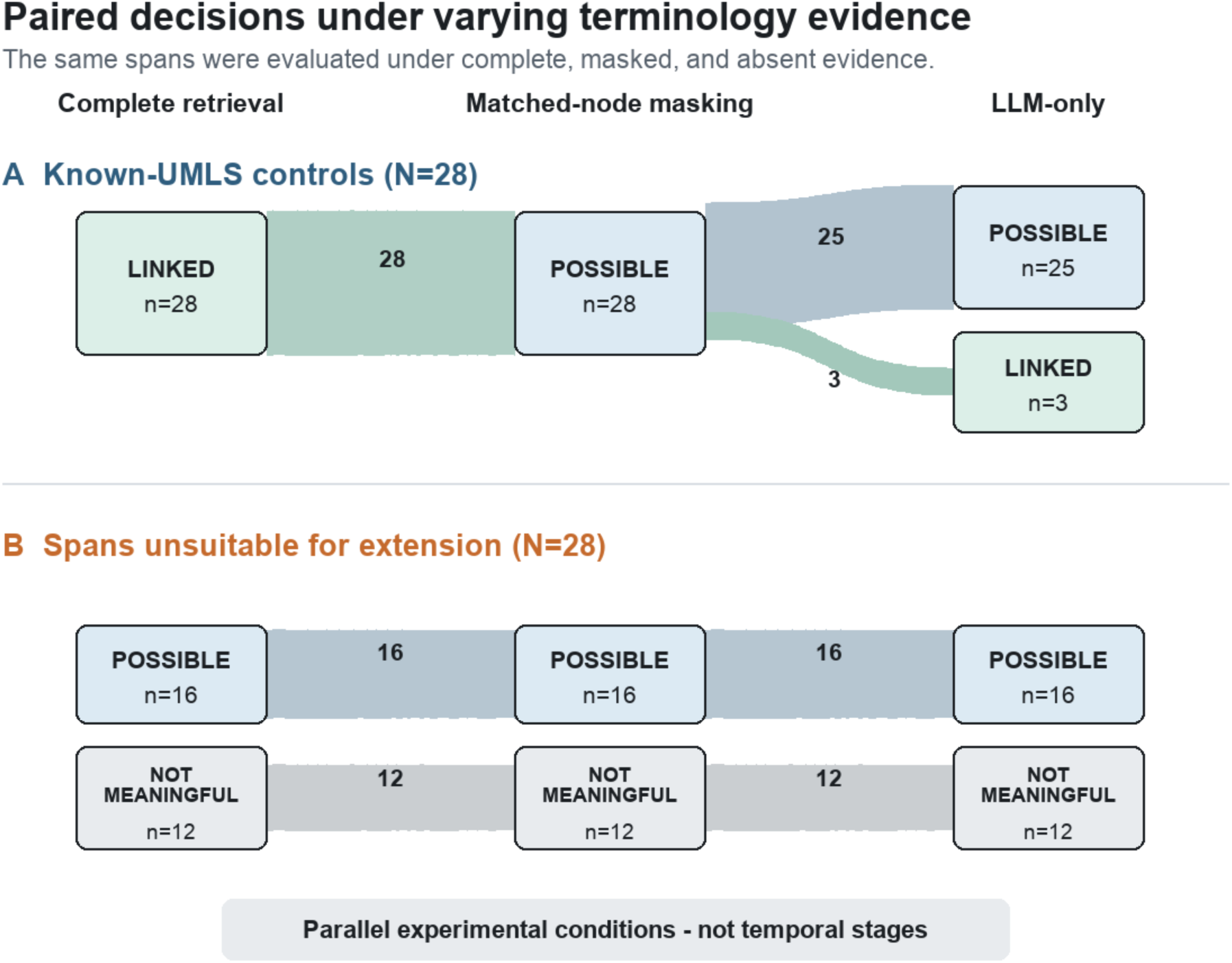
Paired categorical decisions for the same 56 spans under 3 parallel evidence conditions. Among 28 known-UMLS controls, complete retrieval assigned LINKED to all 28, masking assigned POSSIBLE_CANDIDATES to all 28, and LLM-only inference assigned POSSIBLE_CANDIDATES to 25 and LINKED to 3. Among 28 spans adjudicated as unsuitable for ontology extension, the same 12 were labeled NOT_MEANINGFUL and the same 16 were retained as POSSIBLE_CANDIDATES under every condition; none was linked. Band widths are proportional to counts. Horizontal ordering is for display and does not represent a temporal sequence. LLM: large language model; UMLS: Unified Medical Language System.

For the 28 known-UMLS spans, complete retrieval produced a reference-matched link for all 28 (100%; 95% CI, 87.9%-100%). Under matched-node masking, none received an exact link and all 28 were assigned `POSSIBLE_CANDIDATES` (100%; 95% CI, 87.9%-100%). Under LLM-only inference, 25/28 were assigned `POSSIBLE_CANDIDATES` (89.3%; 95% CI, 72.8%-96.3%) and 3/28 were assigned `LINKED` without visible terminology evidence (10.7%; 95% CI, 3.7%-27.2%).

Thus, 25 known spans followed the dominant decision pattern of `LINKED` with complete evidence and `POSSIBLE_CANDIDATES` under both incomplete and absent evidence. The remaining 3 were `LINKED` with complete evidence, assigned `POSSIBLE_CANDIDATES` under matched-node masking, and labeled `LINKED` under LLM-only inference.

Exact McNemar tests yielded *P*<.001 for complete versus masked retrieval and for complete retrieval versus LLM-only inference. Masked retrieval and LLM-only inference differed on 3 items (*P*=.25). Pairwise results and categorical agreement are shown in Table 2.

**Table 2.**
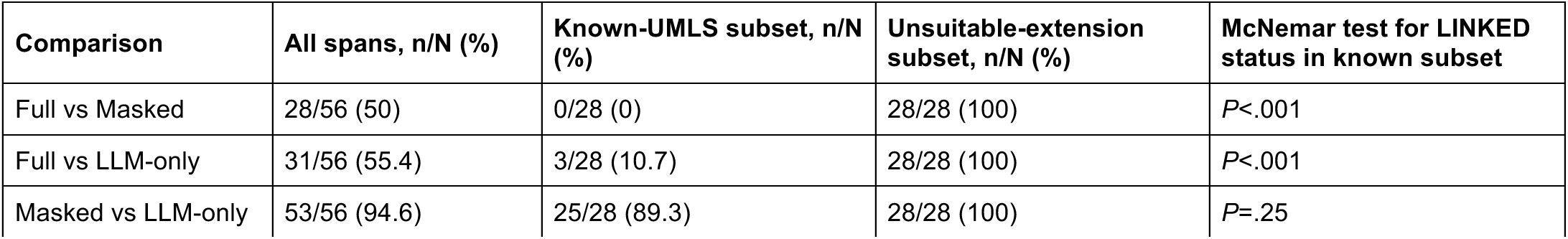
Pairwise exact categorical agreement and exact McNemar tests.

Across all 56 spans, exact categorical agreement was 28/56 (50%) for complete versus masked retrieval, 31/56 (55.4%) for complete retrieval versus LLM-only inference, and 53/56 (94.6%) for masked retrieval versus LLM-only inference. Agreement on the known-UMLS subset was 0/28 (0%), 3/28 (10.7%), and 25/28 (89.3%), respectively. Agreement on the unsuitable-extension subset was 28/28 (100%) for every condition pair.

### Rejection of Spans Unsuitable for Ontology Extension

The 28 spans adjudicated as unsuitable for extension received identical decisions under all 3 evidence conditions. In each condition, 12/28 were classified as `NOT_MEANINGFUL` (42.9%; 95% CI, 26.5%-60.9%) and 16/28 as `POSSIBLE_CANDIDATES` (57.1%; 95% CI, 39.1%-73.5%); none was assigned `LINKED`. The remaining spans were therefore retained for further review rather than assigned exact links.

Reference-concordant rejection rates were 6/7 (85.7%) for MD, 5/11 (45.5%) for OT, 1/4 (25%) for RN, and 0/6 (0%) for SLP (Table 3). These descriptive estimates are imprecise because subgroup sizes were small and uneven; no discipline-level inferential comparisons were performed.

**Table 3.** Reference-concordant rejection of spans unsuitable for ontology extension by note discipline.

| Note discipline | Reference spans, N | Not meaningful, n/N (%) | Possible candidates, n/N (%) | Linked, n/N (%) |
| --- | --- | --- | --- | --- |
| Physician (MD) | 7 | 6/7 (85.7) | 1/7 (14.3) | 0/7 (0) |
| Occupational therapy (OT) | 11 | 5/11 (45.5) | 6/11 (54.5) | 0/11 (0) |
| Nursing (RN) | 4 | 1/4 (25.0) | 3/4 (75.0) | 0/4 (0) |
| Speech-language pathology (SLP) | 6 | 0/6 (0) | 6/6 (100) | 0/6 (0) |
| Overall | 28 | 12/28 (42.9) | 16/28 (57.1) | 0/28 (0) |

### Follow-up Extension-Candidate Study

The follow-up sample contained 84 unique terms, comprising 28 OT, 28 RN, and 28 SLP terms. Each discipline was evenly divided between adult fall-related hospitalization and neonatal intensive care unit strata (14 terms each). All terms were processed with vector retrieval and 10 visible candidates. The top-candidate similarity had a mean of 0.914 (SD 0.027), median of 0.914 (IQR 0.899-0.931), and range of 0.850-0.985.

All 84 terms were classified as `PLAUSIBLE_EXISTING_CONCEPT` (84/84, 100%), and none received a positive extension flag. Table 4 summarizes the sample and retrieval results. Because Study 3 lacked independently adjudicated reference labels for extension candidacy, these findings describe the pipeline’s triage decisions for this initially unmatched sample rather than extension-detection accuracy.

**Table 4.** Follow-up sample composition and full-retrieval results.

| Stratum | Terms, n | Top similarity, mean (SD) | Plausible existing concept, n/N (%) | Extension proposed, n/N (%) |
| --- | --- | --- | --- | --- |
| OT—Falls | 14 | 0.927 (0.028) | 14/14 (100) | 0/14 (0) |
| OT—NICU | 14 | 0.915 (0.024) | 14/14 (100) | 0/14 (0) |
| RN—Falls | 14 | 0.903 (0.027) | 14/14 (100) | 0/14 (0) |
| RN—NICU | 14 | 0.917 (0.030) | 14/14 (100) | 0/14 (0) |
| SLP—Falls | 14 | 0.916 (0.026) | 14/14 (100) | 0/14 (0) |
| SLP—NICU | 14 | 0.909 (0.024) | 14/14 (100) | 0/14 (0) |
| Overall | 84 | 0.914 (0.027) | 84/84 (100) | 0/84 (0) |

### Illustrative Cases From Study 2

“Lumbar spine,” “HTN,” and “pelvis” were linked with complete retrieval, deferred after masking, and linked again by LLM-only inference. For “Lumbar spine,” complete retrieval exposed “Entire lumbar vertebral column,” whereas masked retrieval exposed the alternative candidate “LUMBAR SPINE DISC.” Unsuitable-extension spans retained for review included “grunting and retractions,” “recurrent syncope,” “g-tube placed,” “Return to normal activity when comfortable,” and “functional mobility within room with SBA.” These examples illustrate that evidence-sensitive terminology linking and suitability for ontology extension are distinct judgments.

## Discussion

### Principal Findings

The 3 component evaluations identify distinct sources of variation across a clinical terminology workflow. In the upstream extraction comparison, exact boundary matching produced low F1 for both automated systems and human annotators, whereas embedding-based matching yielded higher agreement and greater separation among the automated systems. Union fusion increased recall at substantial cost to precision, demonstrating that upstream combination rules affect the candidate set presented for terminology linking.

In the paired downstream pilot, exact-link decisions changed with the availability of matched terminology evidence. The decision layer selected the reference concept for all 28 known-UMLS controls when it was visible and deferred all 28 when it was masked. Under LLM-only inference, it deferred 25 controls but assigned `LINKED` to 3 controls without visible terminology evidence.

Because complete retrieval included the reference concept by design, the 100% link rate is a conditional evaluation of the decision layer and does not estimate retrieval recall or routine end-to-end normalization accuracy. Masking likewise represents a controlled retrieval failure rather than the full range of errors encountered in practice. In this benchmark, removing the matched concept produced deferral rather than selection of an alternative retrieved candidate or an extension proposal.

In contrast, the availability of retrieved terminology evidence did not alter decisions for spans adjudicated as unsuitable for ontology extension. All 3 conditions rejected the same 12 spans and retained the same 16. This suggests that exact-link evidence and extension candidacy are distinct decision problems. The first asks whether the visible terminology supports a mapping. The second asks whether an unlinked expression has the abstraction level, compositional form, and expected reuse needed to justify an ontology node.

The follow-up study provides a related observation from an initially unmatched candidate pool. Despite prior screening as absent from UMLS or BioPortal, the pipeline classified all 84 terms as plausible existing concepts after semantic retrieval, and none received a positive extension flag. This finding distinguishes failure to identify an initial lexical or portal match from evidence of genuine concept novelty. However, because independently adjudicated extension labels were unavailable, the correctness of these triage decisions could not be determined.

Taken together, the results illustrate why extraction recall, candidate retrieval, evidence-grounded linkage, and extension candidacy should not be represented by a single end-to-end score. Upstream matching criteria determine which spans appear correct, candidate retrieval determines which terminology evidence becomes visible, and the decision policy determines whether the system links, defers, or rejects. Because the 3 evaluations were not linked at the individual-record level, this interpretation concerns complementary pipeline components rather than a single validated cohort.

### Comparison With Prior Work

Candidate-generation and ranking studies have long shown that downstream normalization is constrained by candidate recall [3–6]. BioSyn and SapBERT improve retrieval representations through biomedical synonym information [4,5], and UMLS-informed generation and ranking provide additional terminology structure [3]. CENT recently demonstrated the benefit of combining semantic candidates with LLM-based disambiguation for clinical procedure normalization [7]. The present ablation complements these accuracy-oriented studies by examining the downstream policy used when decisive candidate evidence is deliberately unavailable.

The results are also consistent with broader RAG evaluation work. Retrieval augmentation can reduce unsupported model content [22], but RAG quality cannot be understood from a single end-to-end score. RAGChecker separates retriever and generator behavior [11], and the RGB benchmark treats negative rejection, noise robustness, and information integration as distinct capabilities [12]. Our complete and masked conditions similarly separate successful evidence use from decision behavior under a controlled retrieval failure.

scispaCy provides robust biomedical pipelines but remains sensitive to domain shift [16], while SciBERT supplies pretrained representations for scientific text [17]. Clinical NER benchmarks have long shown that concept extraction depends on boundary definitions and annotation standards [23]. Nejadgholi et al [24] further showed that exact F1 penalizes some span mismatches that human users may find acceptable, supporting the use of multiple evaluation views rather than a single relaxed score. This study provides a multisite pilot comparison in which the matching definition changed both the absolute agreement level and the apparent separation among extraction systems. The findings are also consistent with recent evidence that LLM-based clinical information extraction can improve on encoder baselines in some settings while introducing practical cost and generalizability tradeoffs [25].

The `POSSIBLE_CANDIDATES` label operationalizes a deferral option analogous to the reject option in selective prediction [14]. Because this study did not assess probabilistic calibration or risk-coverage trade-offs, the label should be interpreted as a categorical workflow decision rather than a calibrated uncertainty measure. CUI-less normalization [15] addresses whether a mention lacks an adequate terminology mapping, whereas ontology-extension candidacy adds a policy judgment about whether the mention should become a reusable concept. The 16 retained unsuitable spans illustrate why these outcomes should not be collapsed.

Biomedical knowledge graph RAG has been used to provide provenance-aware context for model reasoning [9,10]. In terminology linking, provenance is especially important because an exact CUI is a structured claim, not merely a fluent paraphrase. The 3 LLM-only link outputs illustrate the difference between semantic familiarity and evidence for a specific terminology mapping. A familiar expression may invite a plausible answer even when no inspectable candidate supports that answer.

### Implications for Clinical Informatics

Terminology links propagate into downstream data reuse. An unsupported CUI can distort cohort definitions, phenotype extraction, quality measurement, and cross-system interoperability. A conservative deferral policy can reduce that risk, but uniform deferral also creates review workload. Production systems therefore need both high candidate recall and a transparent escalation pathway when evidence is insufficient.

These findings motivate a staged architecture. A first stage should extract candidate spans while retaining confidence, provenance, and alternative boundary interpretations. A second stage should retrieve terminology candidates, establish whether the visible evidence supports an exact match, and attach provenance to any link. If no candidate is adequately supported, the system should expand retrieval or route the span to human review. A third stage should determine extension candidacy using criteria that are not reducible to terminology similarity: compositionality, event or instruction status, granularity, semantic stability, and likely reuse across patients and institutions.

The historical label `NOT_MEANINGFUL` may obscure this distinction because many rejected examples are meaningful in context. A future taxonomy could separate at least 4 states: insufficient retrieval, potentially novel concept, clinically meaningful but compositional content, and documentation fragment or artifact. More precise labels may improve both model learning and adjudicator agreement.

Future evaluation should measure the pipeline at each layer. Extraction measures should report exact, overlap, and application-specific semantic agreement alongside human annotation variability. Retrieval measures should include reference recall at k and coverage across vocabularies. Conditional decision measures should include reference-CUI accuracy when the target is retrieved, unsupported-link rate when it is absent, and deferral rate. Extension triage should be evaluated on independently adjudicated positive and negative candidates. Workflow measures should include reviewer workload, time to resolution, disagreement rates, and the downstream effect of accepted links on semantic data reuse. Structured generation should also be constrained and validated because syntactic conformance does not guarantee evidence-faithful output [26].

### Strengths and Limitations

The study combines a double-annotated, multisite extraction comparison with a paired evidence manipulation and a follow-up extension-triage sample. In Study 2, every span was evaluated with the same prompt and output space under complete, deliberately incomplete, and absent evidence. This design makes the change in exact-link commitment directly interpretable at the decision layer. The inclusion of spans unsuitable for extension also exposes a failure mode that would be hidden by an evaluation limited to known terminology concepts.

The study has several limitations. First, Study 1 included only 66 notes, and model rankings were descriptive without uncertainty intervals or paired comparisons. Exact and embedding-based matching operationalize different notions of agreement; embedding-based scores depend on the representation and matching rule and should not be interpreted as exact clinical correctness. Study 1 used fixed upstream model and semantic-matching configurations, so the observed rankings may not generalize across checkpoints, prompts, similarity thresholds, or matching assignments. BGE-large-en-v1.5 was used both for semantic extraction evaluation and downstream retrieval, introducing dependence on a shared representation model. The 3 evaluations were not linked at the individual-record level and therefore were analyzed separately.

Second, the Study 2 benchmark was small and purpose-built, and the Wilson intervals remain wide. The results should not be interpreted as population-level performance. Third, the known-UMLS reference labels were derived from project adjudication rather than independent gold-standard annotation. Fourth, complete retrieval included the matched concept by design, so its result does not estimate end-to-end retrieval recall. Fifth, masking changed both candidate identity and candidate-set size, preventing separation of matched-node removal from context-volume effects.

Sixth, the Study 2 benchmark contained small and uneven discipline subsets; discipline-specific rejection rates are descriptive only. Seventh, Study 3 was a single, discipline-balanced follow-up sample without independently adjudicated positive and negative reference labels for extension candidacy; it therefore could not estimate extension-detection accuracy. Eighth, Study 2 evaluated a single Gemma 3 27B configuration with default generation settings and a single retriever configuration; robustness across decoding strategies, model checkpoints, and vector-index settings was not assessed. Finally, the findings do not measure clinician outcomes, downstream data quality, or routine review burden.

A larger study should prespecify exact and semantic extraction metrics; incorporate constant-size masking controls, independently adjudicated reference labels, and verified reference CUIs; evaluate repeated runs across multiple retrieval and language models; and include external note sources. It should also include a high-confidence extension-positive set and prospectively define semantic and workflow outcomes.

### Conclusions

Across 3 complementary evaluations analyzed separately, performance depended on both the definition of upstream span agreement and the completeness of downstream terminology evidence. Exact and embedding-based matching produced markedly different NER agreement estimates; complete terminology evidence supported reference-matched linking, whereas removal of the matched node led to consistent deferral. For terms initially unmatched to UMLS or BioPortal, the pipeline classified all 84 as plausible existing concepts after semantic retrieval and proposed none as extensions. These findings support a staged workflow that evaluates extraction, evidence-grounded linking, and extension candidacy separately and treats initial nonmatching as a reason for broader retrieval and review rather than as proof of novelty.

## Data Availability

Aggregate results supporting the reported analyses are presented in the manuscript. The underlying clinical data and term-level records are not publicly available because they originate from protected clinical records and remain governed by institutional privacy controls and data use agreements among the 3 data-contributing institutions. Eligible academic or government investigators may contact the corresponding author, Yibo Chen, PhD, to initiate the institutional request process. Any approved access would occur through the CAIDF secure enclave and would require applicable ethics, privacy, institutional, and data use approvals; access is not guaranteed. Implementation code is not publicly available because it is coupled to institution-specific secure infrastructure and licensed terminology resources.

## Acknowledgments

Services were provided by the University of Iowa’s Institute for Clinical and Translational Science (ICTS); ARPA-H awards to the institute provided direct funding for this project.

YC was affiliated with the MU Institute for Data Science and Informatics at the University of Missouri when the research and analyses reported here were performed and is currently affiliated with the Department of Genetics at Washington University School of Medicine in St. Louis.

The authors thank the CAIDF clinical annotation, terminology, data engineering, and site teams at University of Missouri Health Care, University of Illinois Chicago, and the University of Iowa for creating and stewarding the source corpus and secure research environment.

OpenAI Codex was used during manuscript preparation to assist with grammar correction. It was not used to collect or generate study data or to conduct the original experiments. The authors reviewed and revised all AI-assisted material, independently verified the reported analyses and references, and take full responsibility for the final manuscript.

## Funding

This work was supported by the Creating AI-Enabled All-Health Team Data Fabric (CAIDF) project through an Advanced Research Projects Agency for Health (ARPA-H) award (D24AC00413-00). The content is solely the responsibility of the authors and does not necessarily represent the official views of ARPA-H.

## Data Availability

Aggregate results supporting the reported analyses are presented in this article. The underlying clinical data and term-level records are not publicly available because they originate from protected clinical records and remain governed by institutional privacy controls and data use agreements among the 3 data-contributing institutions. Eligible academic or government investigators may contact the corresponding author, Yibo Chen, PhD, to initiate the institutional request process. Any approved access would occur through the CAIDF secure enclave and would require applicable ethics, privacy, institutional, and data use approvals; access is not guaranteed. Implementation code is not publicly available because it is coupled to institution-specific secure infrastructure and licensed terminology resources.

## Authors’ Contributions

**Proposed wording—YC: conceptualization, methodology, software, formal analysis, investigation, data curation, visualization, writing–original draft, and writing–review and editing. MP: conceptualization, methodology, supervision, project administration, and writing– review and editing.**

## Conflicts of Interest

None declared.

## Abbreviations

CUI: Concept Unique Identifier
LLM: large language model
MD: physician note domain
NER: named entity recognition
NICU: neonatal intensive care unit
OT: occupational therapy
RAG: retrieval-augmented generation
RN: nursing
SLP: speech-language pathology
UMLS: Unified Medical Language System

## Notes

### Competing Interest Statement

The authors have declared no competing interest.

### Author Declarations

This study was a retrospective secondary analysis conducted within the Creating AI-Enabled All-Health Team Data Fabric project. The parent project was reviewed and approved by the University of Illinois Chicago Institutional Review Board under protocol 2024-0553, with required regulatory approvals at the 3 data-contributing institutions. This was not a clinical trial and has no clinical trial registration number. The analyses used deidentified clinical notes and derived terms under institutional data use and governance agreements.

